# Knowledge and Beliefs towards Universal Safety Precautions to flatten the curve during Novel Coronavirus Disease (nCOVID-19) Pandemic among general Public in India: Explorations from a National Perspective

**DOI:** 10.1101/2020.03.31.20047126

**Authors:** Sai Krishna Gudi, Krishna Undela, Rajesh Venkataraman, Uday Venkat Mateti, Manik Chhabra, Sanath Nyamagoud, Komal Krishna Tiwari

**Author notes:** Address for Correspondence: Dr. Sai Krishna Gudi, B.Pharm., PharmD., M.Sc., (Ph.D.), College of Pharmacy, Rady Faculty of Health Sciences, University of Manitoba, Winnipeg, MB, Canada. Authors’ email address: Dr. Krishna Undela, Dr. Rajesh Venkataraman, Dr. Uday Venkat Mateti, Dr. Manik Chhabra, Dr. Sanath Nyamagoud, Dr. Komal Krishna Tiwari.

## Abstract

**Background:** The novel Coronavirus disease (COVID-19) is being considered as the most serious health threat that the world has never witnessed in the recent times and significantly affecting the daily routine of mankind by emerging as a global pandemic. Yet, as there is no treatment nor a vaccine that was approved so far, universal safety precautions (USPs) and mitigating strategies are the only way to deal with this emergency crisis. However, knowledge and beliefs towards USPs among the general public in countries such as India with a large population are lacking.

**Methods:** A prospective, cross-sectional, web-based online survey was conducted among the general public in India during March 2020. A 20-item self-administered survey questionnaire was developed and randomly distributed among the public using google document forms through social media networks. Descriptive statistics were used in representing the study characteristics, and the Chi-square test was used in assessing the associations among the study variables with a p-value of < 0.05 was considered as statistically significant.

**Results:** Of 1287 participants, 1117 have given their consent of willingness and completed the questionnaire with a response rate of 86.8%. The mean age of the study participants was 28.8 ± 10.9 years, where the majority of them belong to the age category <25 years, and sex was equally distributed. Based upon the socio-demographic information, the majority were post-graduates (32.9%), professional job holders (45%) and belonged to the upper-middle (40%) economic class. Overall, the knowledge and beliefs towards USPs and mitigating strategies among participants varied between moderate to high, with statistically significant associations with their socio-demographic characteristics.

**Conclusions:** Although the knowledge and beliefs of the general public in India towards USPs are encouraging, there is a need for long-term educational interventions as the dynamics and severity of COVID-19 have been changing day-by-day rapidly. The findings of this study could guide the public health authorities in making and implementing decisions to combat this pandemic.

## Introduction

Novel coronavirus (SARS-CoV-2), formally named as the novel Coronavirus-2019 (nCoV-2019) by the World Health Organization (WHO) after identifying a cluster of pneumonia cases during late December 2019 in the province of Hubei, China [1]. However, coronavirus (CoV) is not the first-ever occurrence as it was initially discovered in the 1960s and belongs to a family *coronaviridae* [2]. Since then, several types of CoV were identified amongst which Severe Acute Respiratory Syndrome-CoV (SARS-CoV) and Middle East Respiratory Syndrome-CoV (MERS-CoV) are of importance [3]. As of 27 March 2020, a total of 509,164 confirmed cases and 23,335 deaths of Coronavirus disease (COVID-19) have been reported globally, effecting 151 countries, where around half of those reported cases and deaths are solely attributed to China [4]. However, within a short span of time, the cumulative proportion of cases and deaths outside China has been outstripped by the total number of cases and deaths in China, which has raised significant concern among the public around the globe. As a result, considering the rapid disease transmission of COVID-19 within and across the countries, on 11 March 2020, WHO has declared the COVID-19 outbreak as a global pandemic [5].

Although initial investigations reported that the spread of COVID-19 would be possibly from animals-to-humans, later investigations have stated that human-to-human transmission also could occur [6]. At this initial stage, the definite modes of this pandemic are not completely known; however, health officials suggest that it could primarily spread through the droplets when an infected person coughs or sneezes and by direct contact with the infected individuals [7]. Unfortunately, no drug has been officially approved for the treatment of this global pandemic, although drugs such as hydroxychloroquine and remdesivir are under clinical investigation [8]. In this manner, acquiring and being adherent to the universal safety precautions (USP) is the only choice in controlling the widespread of COVID-19 across the world.

Maintaining personal hygiene is an essential practice to be protected against any type of respiratory illness, such as COVID-19. Of this note, hand washing has been considered as an effective measure in preventing the cross-transmission from one to another [9]. Social distancing is a technique where actions are taken to reduce the frequency of contacts and to maintain a proper distance between individuals in order to limit the transmission of any communicable disease [10]. Usually, social distancing would be in-place where the spread has been believed to happen in a broader community. Other social distancing measures include isolation and quarantine, which also play a major role, particularly among those individuals presenting with symptoms, including confirmed cases. Isolation is an act of separating the ill-persons with a contagious disease from a non-infected person to reduce the spread [11]. Usually, isolation procedures are commonly observed in hospital settings. While, quarantine is considered as one of the effective methods in controlling communicable outbreaks and pandemics such as COVID-19, at a community level, where the movement of a person is restricted to the home or a designated facility, who are presumed to have been exposed to a contagious disease, but without having any symptoms [12].

If all the above detailed social distancing measures are insufficient in reducing the widespread of the infection, community containment would be implemented where an entire community or neighbourhood is restricted to reduce personal interactions, except for inevitable situations. Besides, usage of personal protective equipment (PPE) would be another precautionary measure to be taken in controlling further spread during outbreaks, although PPE is considered as less effective compared to other control measures. Routinely, PPEs are widely used in healthcare settings as a standard to protect healthcare workers from infections [13]. Among different types of PPEs, face masks and hand gloves are the most commonly used in situations during emergency outbreaks, especially by the sick people in order to avoid the spread. Face masks have always been popular and well-known public interventions used as a self-protection measure. The use of face masks has been included in their pandemic plans by most of the developed countries such as the United States, France and Australia [14]. WHO states that wearing a face mask incorrectly may actually increase the chance of being infected, rather than decreasing [15]. Thus, the correct use of face masks is particularly essential while dealing with the situations during pandemics such as COVID-19. Implementing the above-mentioned non-pharmacological USPs during the COVID-19 pandemic would relieve the concerns of over-loaded healthcare systems as well as the public to a remarkable extent.

Although fewer studies have attempted in assessing the knowledge aspects related to the COVID-19 among the healthcare workers, authors of this study strongly believe that focusing on the USPs such as, maintaining personal hygiene, washing hands, adhering to social-distancing techniques, self-isolation practices, usage of face masks and hand sanitizers would significantly contribute in lowering the spread and eventually flatten the curve. So far, the course of the COVID-19 in India seems under control with lower rates of incidence despite the large population. However, looking at the dynamics of SARS-CoV-2, especially its higher rates of transmission, things could alter unpredictably, leading to adverse situations, at any time. Of concern in this regard, examining the knowledge and beliefs regarding USPs and mitigating strategies among the general public in India during this pandemic would be a chief priority.

## Methods

This is a prospective, cross-sectional, and web-based online survey conducted using a questionnaire with an intention to obtain the responses regarding USPs and mitigating strategies towards COVID-19 among the general public in India during March 2020.

### Development & Content of Survey Instrument

A 20-items, semi-structured and self-administered questionnaire was developed using factsheets, course materials, information leaflets and booklets developed by Health Protection Surveillance Centre (HPSC) [16], Public Health Agency (HSC) [17], Centre for Disease Control and Prevention (CDC) [18], National Health Service (NHS) [19], and WHO [20] for the purpose of COVID-19 prevention. The study questionnaire has two domains, i.e., knowledge and belief domain, which have ten questions each. Of particular interest towards preventive measures, questions in both the domains have focussed on aspects such as personal hygiene, proper handwashing, use of face masks, and social distancing techniques such as quarantine and isolation.

### Reliability & Validity of Survey Instrument

Firstly, the developed questionnaire was validated using face and content validation methods by the selected faculty members and researchers to ensure the readability. Secondly, it was assessed for reliability using test-retest and split-half methods through a pilot study that was pre-tested among 30 random respondents for the clarity, acceptability and relevance, prior to the full-fledged survey. Lastly, the survey questionnaire was distributed among the public after revising the comments and considering the pre-test results to facilitate better comprehension.

### Distribution of the Survey Questionnaire

Following the reliability and validity assessments, the survey questionnaire was distributed across the social media and professional platforms such as WhatsApp, Facebook, Messenger, Gmail, Twitter, LinkedIn, Outlook and Telegram using the Google forms [21].

### Sampling method

With the fact that this is an online web-based survey which is exploratory in nature, a non-probability sampling method with an invitation online sampling technique was used.

### Ethical Considerations

Participants were explained about the purpose of the study and requested to provide the consent of voluntary willingness before participating in the survey. All the procedures performed in this study involving human participants were in adherence to the ethics of the 1964 Helsinki declaration and its later amendments or comparable ethical standards. This study was conducted and reported according to the Checklist for Reporting Results of Internet E-Surveys guidelines (CHERRIES) [22].

### Statistical Analysis

Data were entered into Microsoft Excel spreadsheets and cross-checked for accuracy and the statistical analysis was performed using IBM SPSS software for windows, version 24 (Armonk, NY, USA) [23]. Descriptive statistics on sample characteristics were computed, including means, standard deviation, and frequency distributions. Variables included in the analysis were age, sex, educational level, occupation and economic status (as per modified Kuppuswamy scale) [24]. The Chi-square test was used in assessing the associations among the study variables and a p-value of < 0.05 was considered as statistically significant.

## Results

Of 1287 participants that filled out the survey, a total of 1117 participants have given their consent of willingness and completed the questionnaire with a response rate of 86.8%. The mean age of the study participants was 28.8 ± 10.9 years, where more than half of the participants belong to the age category <25 years. Sex was equally distributed among the participants, while more than 90% of them had either doctoral (28.4%), post-graduate (32.9%) or under-graduate (30.1%) degree as their educational background. The majority of the participants were professional job holders (45%) and then followed by the students (32%), while around 40% of the study participants belong to the upper-middle economic class. All other socio-demographic characteristics are detailed in Table 1.

**Table 1:** Sociodemographic characteristics of study participants (N=1117)

| **Variables** | **Participants (N)** | **Percentage (%)** |
| --- | --- | --- |
| **Age** |  |  |
| *Mean age ± S.D* $28.78 \pm 10.88$ | | |
| < 25 years | 666 | 59.6 % |
| 25-45 years | 284 | 25.4% |
| > 45 years | 167 | 15.0% |
| **Sex** |  |  |
| Male | 568 | 50.9% |
| Female | 549 | 49.1% |
| **Level of education** |  |  |
| Doctoral degree | 317 | 28.4% |
| Post-graduate | 367 | 32.9% |
| Under-graduate | 336 | 30.1% |
| Post-secondary/Diploma | 38 | 3.4% |
| Secondary education | 48 | 4.3% |
| High school | 11 | 1.0% |
| **Occupation status** |  |  |
| Professional | 503 | 45.0% |
| Skilled worker | 124 | 11.1% |
| Unskilled worker | 46 | 4.1% |
| Unemployed | 13 | 1.2% |
| Retired employee | 53 | 4.7% |
| Homemaker | 21 | 1.9% |
| Student | 357 | 32.0% |
| **Economical class (rupees/month)** |  |  |
| Upper ($\geq 52,734$) | 272 | 24.4% |
| Upper middle (26,355-52,733) | 445 | 39.8% |
| Lower middle (19,759-26354) | 45 | 4.0% |
| Upper lower (13,161-19,758) | 295 | 26.4% |
| Lower ($\leq 13,160$) | 60 | 5.4% |

### Knowledge towards universal safety precautions

Although more than two-thirds of the study participants provided the correct response, the rest of the 30% were not aware of the ideal length of time to wash hands in preventing the infection. Surprisingly, around half of the participants were not aware of the ideal strength of the alcohol that a hand-sanitizer should contain to be used during outbreaks, while more than two-thirds of the participants were not known about the ideal distance to be maintained as part of social distancing measures. Around three-fourth and two-fifth of the participants were not aware of the quarantine and isolation procedures that are usually followed during outbreaks. However, the majority of the participants (i.e., 84%, 77% & 94%) are knowledgeable about the self-isolation time period, type of the face mask to be used, and social distancing procedures, respectively. The rest of the information about the knowledge of the general public towards USPs are detailed in Table 2. Furthermore, correct responses for the respective questions with their 95% confidence intervals are represented in Figure 1.

**Table 2:** Knowledge towards universal safety precautions among study participants.

| Questions | Correct answer | Correct Response N (%) | Incorrect Response N (%) |
| --- | --- | --- | --- |
| 1. What would be the ideal length of time to wash hands in preventing the spread of COVID-19? | 20 seconds | 779 (69.7%) | 338 (30.3%) |
| 2. What is the ideal strength of alcohol that a hand-sanitizer should contain to be used during outbreaks? | 60 % | 603 (54.0%) | 514 (46.0%) |
| 3. What would be the ideal distance to be maintained in preventing the spread of COVID-19? | 2 meters/6 feet | 338 (30.3%) | 779 (69.7%) |
| **4.** During outbreaks, ‘*quarantine*’ is a procedure usually followed by? | People who are at risk | 286<br>(25.6%) | 831<br>(74.4%) |
| **5.** During outbreaks, ‘*isolation*’ is a procedure usually followed by? | Infected people | 654<br>(58.5%) | 463<br>(41.5%) |
| **6.** Health status of all the close contacts should be monitored for how many days from the last exposure (i.e., *self-isolation period*)? | 14 days | 940<br>(84.2%) | 177<br>(15.8%) |
| **7.** Which type of face mask is considered as the ideally protective during outbreaks? | N-95 face mask | 857<br>(76.7%) | 260<br>(23.3%) |
| **8.** *Social distancing* means staying away from the crowd and maintaining minimum distance from people around, with the intention of minimizing the transmission of an outbreak. | True | 1045<br>(93.6%) | 72 (6.4%) |
| **9.** The term ‘*close contacts*’ are those who had provided care for infected persons? | True | 705<br>(63.1%) | 412<br>(36.9%) |
| **10.** The term ‘*transient contacts*’ are those who had interacted with the infected persons for a short period of time? | True | 821<br>(73.5%) | 296<br>(26.5%) |

**Figure 1:**
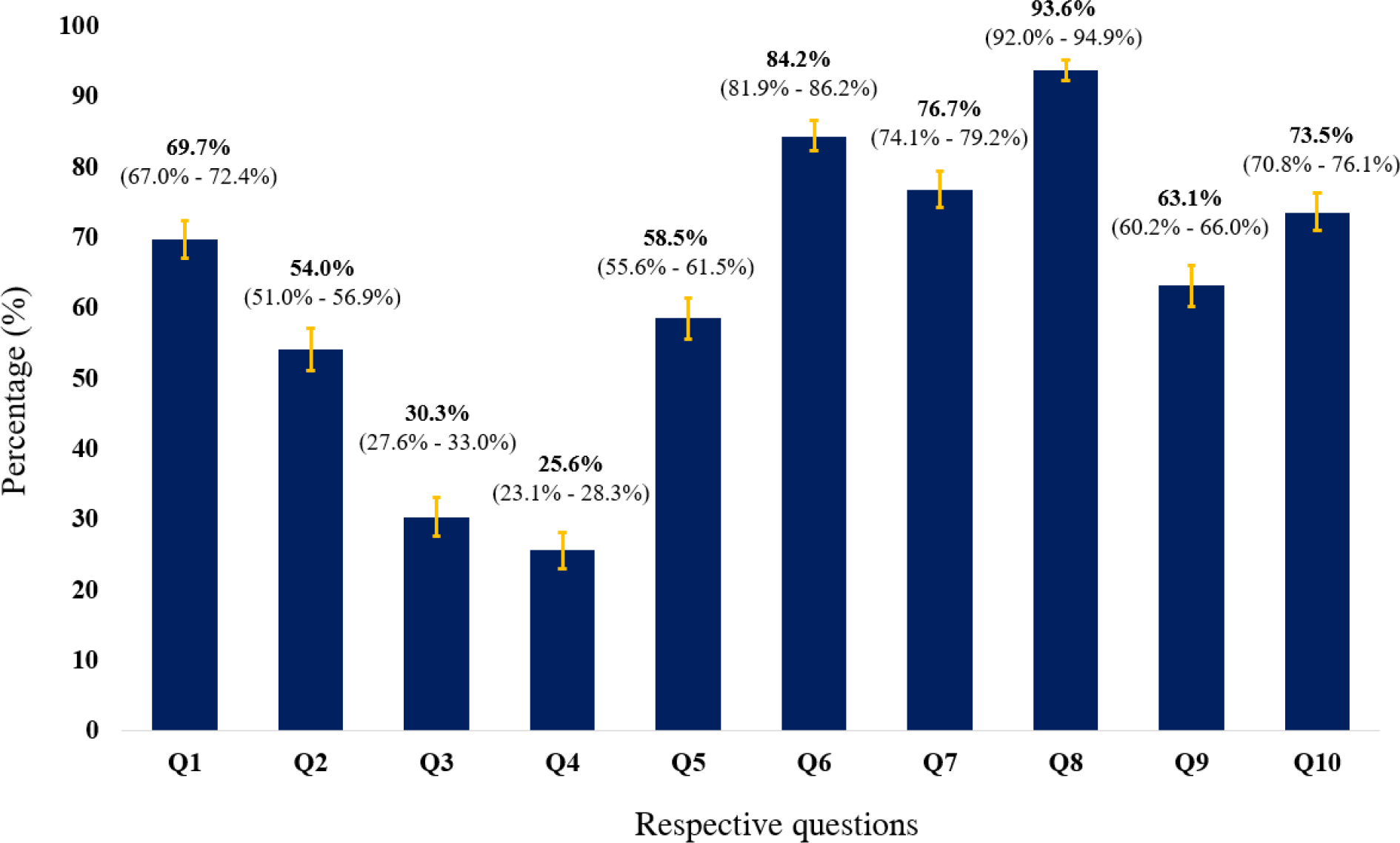
Proportions of correct responses for questions in the knowledge domain with 95% CI

### Beliefs towards universal safety precautions

The majority of the study participants (94%) believe that maintaining good personal hygiene, washing hands frequently using soap, avoiding handshaking behaviour and avoiding placing fingers in eyes, nose and mouth would prevent the spread of the infection. While more than 96% of the participants believe that adhering to social distancing measures and staying at home would be a good practice in combating the spread. However, notably, three-fourth of the participants (75%) believe that wearing a face mask is considered appropriate and protective even in the absence of the symptoms. The rest of the information about the beliefs of the general public towards USPs are detailed in Table 3. The correct responses for the respective questions with their 95% confidence intervals are represented in Figure 2. Furthermore, the association of socio-demographic variables with the general public’s beliefs were assessed using the chi-square test and presented in Table 4.

**Table 3:** Beliefs towards universal safety precautions among study participants

| Questions | Correct answer | Correct Response N (%) | Incorrect Response N (%) |
| --- | --- | --- | --- |
| 1. Maintaining good personal hygiene and being socially responsible would prevent the spread of COVID-19? | Agree | 1045 (93.6%) | 72 (6.4%) |
| 2. Washing hands frequently using soap or sanitizer would prevent the spread of COVID-19? | Agree | 1045 (93.6%) | 72 (6.4%) |
| 3. Avoiding handshaking behaviour would prevent the spread of COVID-19? | Agree | 1047 (93.7%) | 70 (6.3%) |
| 4. Avoiding placing fingers into eyes, nose and mouth would prevent the spread of COVID-19? | Agree | 1050 (94.0%) | 67 (6.0%) |
| **5.** Coughing and sneezing into the elbow or within the clothing is a good practice in preventing the spread of COVID-19? | Agree | 961<br>(86.0%) | 156<br>(14.0%) |
| **6.** Limiting eating and sharing food with colleagues and friends would prevent the spread of COVID-19? | Agree | 745<br>(66.7%) | 372<br>(33.3%) |
| **7.** Following social distancing measures and avoiding crowded places would limit the spread of COVID-19? | Agree | 1076<br>(96.3%) | 41 (3.7%) |
| **8.** Wearing a face mask is considered appropriate and protective, although not having any symptoms? | Disagree | 283<br>(25.3%) | 834<br>(74.7%) |
| **9.** Proper usage of face mask should include covering nose, mouth and chin with the coloured side facing outside? | Agree | 978<br>(87.6%) | 139<br>(12.4%) |
| **10.** Staying at home would play a significant role in preventing the spread of COVID-19? | Agree | 1077<br>(96.4%) | 40 (3.6%) |

**Table 4:** Association between socio-demographic variables and the general public’s beliefs towards USPs

| Qu<br>esti<br>ons | Educational Level |  |  |  |  |  |  |  | Occupational Status |  |  |  |  |  |  |  | Economic Class |  |  |  |  |  |
| --- | --- | --- | --- | --- | --- | --- | --- | --- | --- | --- | --- | --- | --- | --- | --- | --- | --- | --- | --- | --- | --- | --- |
|  | Options | 1 | 2 | 3 | 4 | 5 | 6 | p-value | 1 | 2 | 3 | 4 | 5 | 6 | 7 | p-value | 1 | 2 | 3 | 4 | 5 | p-value |
| Q1 | Not Sure | 2 | 2 | 9 | 9 | 11 | 0 | <0.001 | 7 | 12 | 9 | 0 | 2 | 3 | 0 | <0.001 | 3 | 17 | 5 | 8 | 0 | <0.001 |
|  | Agree | 304 | 359 | 316 | 29 | 28 | 9 |  | 484 | 105 | 33 | 21 | 344 | 45 | 13 |  | 267 | 413 | 37 | 270 | 58 |  |
|  | Disagree | 11 | 6 | 11 | 0 | 9 | 2 |  | 12 | 7 | 4 | 0 | 11 | 5 | 0 |  | 2 | 15 | 3 | 17 | 2 |  |
| Q2 | Not Sure | 6 | 0 | 5 | 9 | 6 | 2 | <0.001 | 7 | 6 | 9 | 2 | 4 | 0 | 0 | <0.001 | 5 | 13 | 2 | 6 | 2 | 0.000 |
|  | Agree | 303 | 354 | 316 | 29 | 34 | 9 |  | 473 | 111 | 35 | 19 | 347 | 47 | 13 |  | 259 | 413 | 37 | 278 | 58 |  |
|  | Disagree | 8 | 13 | 15 | 0 | 8 | 0 |  | 23 | 7 | 2 | 0 | 6 | 6 | 0 |  | 8 | 19 | 6 | 11 | 0 |  |
| Q3 | Not Sure | 2 | 5 | 5 | 12 | 8 | 0 | <0.001 | 13 | 6 | 9 | 0 | 4 | 0 | 0 | <0.001 | 7 | 13 | 2 | 10 | 0 | 0.001 |
|  | Agree | 303 | 356 | 316 | 26 | 35 | 11 |  | 478 | 111 | 35 | 21 | 339 | 50 | 13 |  | 261 | 418 | 36 | 274 | 58 |  |
|  | Disagree | 12 | 6 | 15 | 0 | 5 | 0 |  | 12 | 7 | 2 | 0 | 14 | 3 | 0 |  | 4 | 14 | 7 | 11 | 2 |  |
| Q4 | Not Sure | 2 | 3 | 11 | 12 | 7 | 0 | <0.001 | 14 | 6 | 9 | 0 | 4 | 2 | 0 | <0.001 | 12 | 11 | 2 | 10 | 0 | 0.316 |
|  | Agree | 306 | 360 | 315 | 22 | 36 | 11 |  | 483 | 111 | 33 | 21 | 344 | 45 | 13 |  | 254 | 421 | 40 | 275 | 60 |  |
|  | Disagree | 9 | 4 | 10 | 4 | 5 | 0 |  | 6 | 7 | 4 | 0 | 9 | 6 | 0 |  | 6 | 13 | 3 | 10 | 0 |  |
| Q5 | Not Sure | 2 | 5 | 19 | 10 | 9 | 0 | <0.001 | 7 | 8 | 11 | 0 | 16 | 3 | 0 | <0.001 | 8 | 20 | 7 | 8 | 2 | 0.05 |
|  | Agree | 288 | 331 | 275 | 23 | 35 | 9 |  | 455 | 103 | 32 | 16 | 302 | 44 | 9 |  | 239 | 384 | 31 | 253 | 54 |  |
|  | Disagree | 27 | 31 | 42 | 5 | 4 | 2 |  | 41 | 13 | 3 | 5 | 39 | 6 | 4 |  | 25 | 41 | 7 | 34 | 4 |  |
| Q6 | Not Sure | 27 | 31 | 24 | 11 | 10 | 3 | <0.001 | 40 | 22 | 15 | 0 | 29 | 0 | 0 | <0.001 | 23 | 50 | 6 | 25 | 2 | 0.244 |
|  | Agree | 223 | 253 | 219 | 22 | 26 | 2 |  | 357 | 67 | 24 | 12 | 240 | 39 | 6 |  | 190 | 292 | 25 | 192 | 46 |  |
|  | Disagree | 67 | 83 | 93 | 5 | 12 | 6 |  | 106 | 35 | 7 | 9 | 88 | 14 | 7 |  | 59 | 103 | 14 | 78 | 12 |  |
| Q7 | Not Sure | 4 | 0 | 5 | 9 | 6 | 0 | <0.001 | 7 | 6 | 9 | 0 | 2 | 0 | 0 | <0.001 | 5 | 9 | 2 | 8 | 0 | <0.001 |
|  | Agree | 311 | 363 | 326 | 29 | 36 | 11 |  | 494 | 113 | 37 | 21 | 351 | 47 | 13 |  | 267 | 430 | 37 | 282 | 60 |  |
|  | Disagree | 2 | 4 | 5 | 0 | 6 | 0 |  | 2 | 5 | 0 | 0 | 4 | 6 | 0 |  | 0 | 6 | 6 | 5 | 0 |  |
| Q8 | Not Sure | 10 | 22 | 14 | 11 | 8 | 2 | **<0.001** | 31 | 6 | 9 | 2 | 17 | 2 | 0 | **0.005** | 11 | 29 | 4 | 21 | 2 | 0.469 |
|  | Agree | 227 | 252 | 235 | 18 | 28 | 7 |  | 353 | 84 | 29 | 12 | 251 | 32 | 6 |  | 185 | 303 | 30 | 201 | 48 |  |
|  | **Disagree** | 80 | 93 | 87 | 9 | 12 | 2 |  | 119 | 34 | 8 | 7 | 89 | 19 | 7 |  | 76 | 113 | 11 | 73 | 10 |  |
| Q9 | Not Sure | 24 | 21 | 30 | 14 | 9 | 0 | **<0.001** | 32 | 23 | 13 | 5 | 18 | 7 | 0 | **<0.001** | 15 | 42 | 13 | 24 | 4 | **<0.001** |
|  | **Agree** | 284 | 335 | 292 | 24 | 32 | 11 |  | 455 | 98 | 29 | 16 | 326 | 43 | 11 |  | 251 | 389 | 27 | 257 | 54 |  |
|  | Disagree | 9 | 11 | 14 | 0 | 7 | 0 |  | 16 | 3 | 4 | 0 | 13 | 3 | 2 |  | 6 | 14 | 5 | 14 | 2 |  |
| Q10 | Not Sure | 4 | 5 | 3 | 9 | 9 | 0 | **<0.001** | 8 | 6 | 9 | 0 | 4 | 3 | 0 | **<0.001** | 4 | 9 | 5 | 12 | 0 | **<0.001** |
|  | **Agree** | 313 | 362 | 326 | 29 | 36 | 11 |  | 491 | 115 | 37 | 21 | 353 | 47 | 13 |  | 268 | 432 | 37 | 280 | 60 |  |
|  | Disagree | 0 | 0 | 7 | 0 | 3 | 0 |  | 4 | 3 | 0 | 0 | 0 | 3 | 0 |  | 0 | 4 | 3 | 3 | 0 |  |

*All the correct responses and the statistically significant p-values (< 0.05) were bolded.*
*Where, educational Level (1-Doctoral degree, 2-Post-graduate, 3-Under-graduate, 4-Post-secondary/Diploma, 5-Secondary education, 6-High school); occupation status (1-Professional, 2-Skilled worker, 3-Unskilled worker, 4-Unemployed, 5-Retired employee, 6-Home maker, 7-Student); economic class (1-Upper, 2-Upper middle, 3-Lower middle, 4-Upper lower, 5-Lower.*

**Figure 2:**
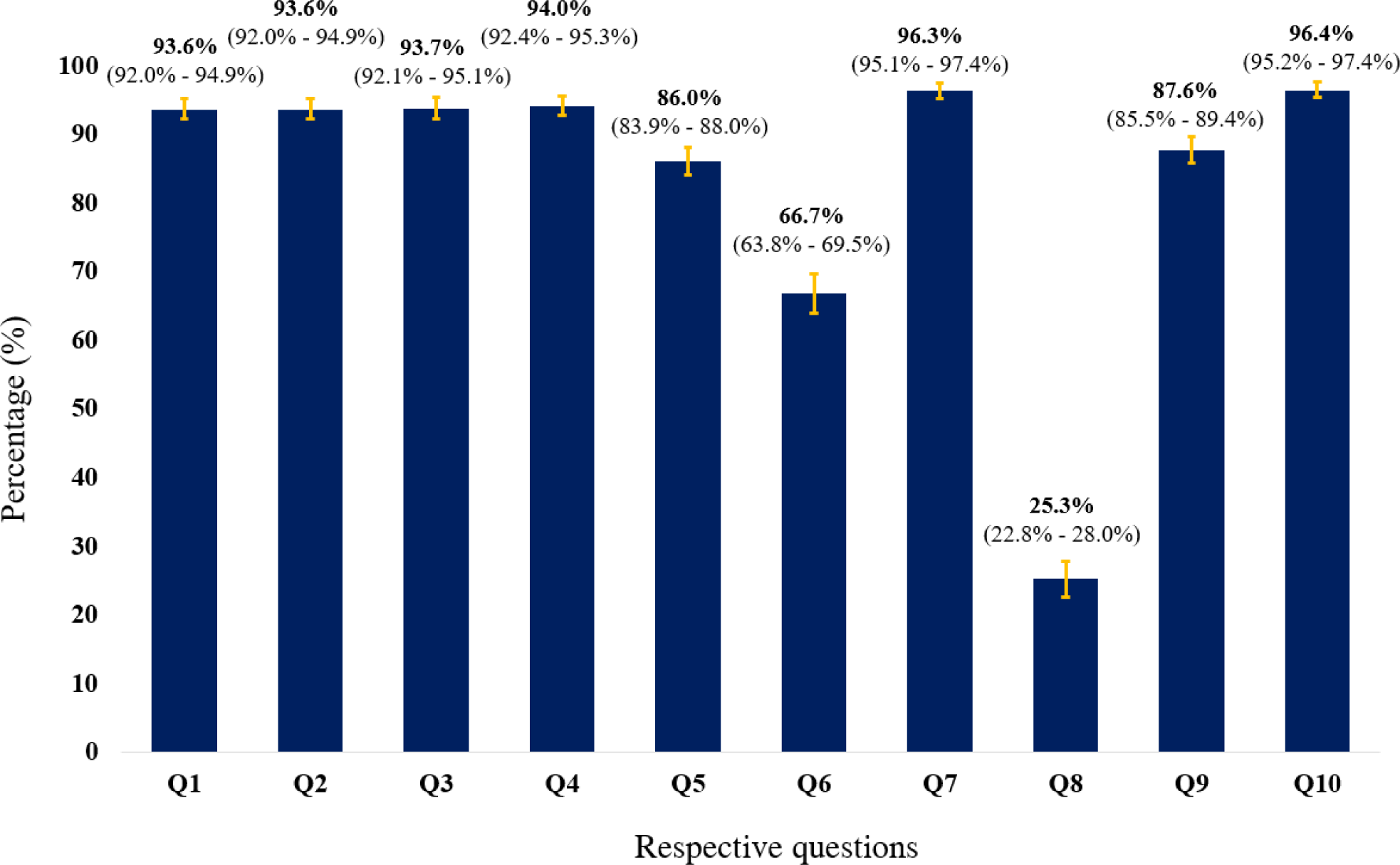
Proportions of correct responses for questions in beliefs domain with 95% CI

## Discussion

This current study is believed to be the first of its kind among the general public in India to assess the knowledge and beliefs towards USPs in combating the spread of COVID-19. Our study results noticed that although the knowledge of the participants regarding the self-isolation time periods (84%) and the social distancing measures (94%) was relatively high, knowledge regarding the ideal length of time to wash hands (70%), the strength of alcohol that a hand-sanitizer should contain to be used during outbreaks (54%), ideal distance to be maintained in preventing the spread of COVID-19 (30%), quarantine and isolation procedures (26% & 58%), type of face mask to be used (77%), close and transient contacts (63% & 73%) were found to be insufficient. Overall, correct responses for the beliefs towards USPs among study participants were found be relatively high, except for the questions regarding eating and sharing food with colleagues and friends during COVID-19 pandemic (67%) and wearing a face mask without having any symptoms (25%), which informs the need for educational interventions among the general public in India.

Within a no time, COVID-19 has evolved as a world pandemic after declaring it as an epidemic and thus considering the current situations with no approved treatment yet, USPs and community mitigation strategies, such as maintaining personal hygiene, handwashing behaviour, social distancing, isolation & quarantine techniques, rational usage of face masks could play a major role in diminishing the virus transmission to a significant extent. Availing today’s technology, certain steps such as sending alerts, disease updates, figures & facts, and precautionary measures regarding the outbreak through automatic text messages or pop-ups should be sent out by the respective telecommunicators in informing the public. An exploratory pilot study conducted in the United Kingdom that looked at the development of interventions in reducing the respiratory infections and flu, which also looked at the measurement and prediction of handwashing intentions, concluded that sending high-threat and coping messages to the public would increase their handwashing intentions [25]. However, at times, threat messages would be ineffective and might also lead to counter-productive effects that have to be kept in mind.

Generally, educating the public with an emphasis on personal hygiene, such as washing hands, would take a lot of time and effort, especially in developing countries such as India. In a large cross-sectional comparative study conducted in Bangladesh from 2006 to 2011, including participants from 50 sub-districts inferred that there exists a significant gap between perceptions and practice of proper handwashing behaviours among their study participants. It also found that handwashing behaviour before eating food was lower, and unfortunately, only 8% of their study participants stated that they use soap for washing their hands at the baseline. It also noticed that handwashing knowledge and practices were relatively lower before cooking, serving and eating food [26]. Furthermore, socioeconomic status, including education, have shown a positive association with handwashing, which are similar to our current study findings. During outbreaks such as COVID-19, information regarding safety measures should be selflessly promoted by the news channels, print media, radio stations, and social media, as almost every individual relates to either of these platforms at some point in a day. Around three fourth of respondents in a descriptive cross-sectional study conducted in Nigeria have stated that they have acquired good handwashing measures by watching health education messages from social media, newspapers and radio channels [27].

In contrast with the current study findings, a study conducted during an outbreak of H1N1 influenza A pandemic in 2009 at a large public university has found poor compliance with the mitigation strategies such as staying at home while ill in avoiding the spread of the virus. In addition, around half of their study participants, including students, staff and faculty members, had attended the social gatherings despite acute respiratory infection and feeling ill [28]. Besides government and healthcare organizations, efforts should be made by the local researchers in conducting awareness and education camps by actively taking part in participatory, community-based research, which could have a remarkable impact on the safety practices among the public. A systematic review that looked at identifying the handwashing techniques in primary and community levels concluded that only a few studies have demonstrated the proper handwashing techniques and noticed that there is a lack of evidence for handwashing techniques are being used in practice today. Moreover, it also found that most of the studies that assessed the handwashing measures were poorly designed along with the inconclusive statements [29].

Among various infection control strategies, usage of personal protective equipment (PPE) is necessarily essential during outbreaks such as COVID-19 among healthcare workers and infected people with an intention to prevent the transmission of the disease. Although PPEs are not recommended as the front-line defence measures, these should be in place along with the other administrative and environmental control measures [30]. PPEs such as face masks, respirators, hand gloves, goggles and face shields are of the utmost importance when the outbreak or pandemic is in its early stage, especially when there are no vaccines or medications available.

Unfortunately, a recent systematic review that looked at the use of PPEs to protect against respiratory infections in Pakistan has inferred that PPEs were not available at many facilities, and its use was limited during high-risk situations [30]. Furthermore, it observed lower compliance towards PPEs among healthcare workers with reuse of PPEs behaviour. Respective countries should implement stringent policies to avoid such practices, which could potentially lead to the spread of the infection. In a cross-sectional survey that evaluated the knowledge, use and barriers towards the PPEs for airway management among emergency medical technicians during SARS outbreak in Canada have reported that most of their study participants opined that N-95 respirator mask is the safest and should be used during outbreaks which are in-line with the current study findings [13]. However, the appropriate selection and efficacy of the respiratory protective apparatus have always been a controversy. As these coronaviruses are extremely small, they will have the ability to pass through the pores of both the surgical mask and the N-95 mask. Although the N-95 respirator apparatus has a good structure, which prevents the passage of virus, if it is contaminated and reused, there are high chances for it to become the source of infection instead [31]. The current study findings related to the proper use of a face mask are in accordance with a cross-sectional study that assessed the usage of face masks in a primary care outpatient setting in Hong Kong, where more than half of their participants have demonstrated the correct steps in wearing a face mask [32].

Although usage of face masks itself might not be an effective intervention in combating the spread of infection, it could be used as an adjunct measure, especially among those who are ill and infected patients. Adopting the use of face masks during outbreaks could be affected by various factors such as social acceptability, perception of the disease risk, need for the mask, and comfort & fit of the mask [33]. Interestingly, a Cochrane intervention review by Verbeek JH et al. concluded that usage of PPEs, especially face masks, might not lead to more containment but may have more user satisfaction [34].

On the flip side, implementing mitigation strategies such as isolation, quarantine and community containment during outbreaks would have major challenges, which includes early case detection in order to self-isolate, need for psychological support and availability of basic needs while being separated from the public, and the most importantly, dealing and managing with the ethical codes, principles, self-determination, individual conflicts and rights of liberty of the public are major challenges that should be considered while implementing such strategies during pandemics such as COVID-19 [11]. Furthermore, a systematic review that critically evaluated the role and impact of social distancing measures during influenza pandemic had attempted various mitigating interventions such as school & workplace closure, home working, self-isolation, quarantine of contacts, mobility restriction, and cancellation of mass events along with their effectiveness [35]. Firstly, it observed that either proactive or reactive school closure found to be moderately effective in reducing the transmission of influenza and could only delay the peak of the pandemic by a week; however, it was associated with the high secondary cost. It also mentioned that the main purpose of the school closure policy during the pandemic might not be optimally achieved as most of the children may engage in outdoor activities during the school closure period. Secondly, a similar type of results was noticed with the workplace closure and home working interventions, where they are modestly effective and acceptable, while increases the secondary cost. Instead, these interventions would affect workplace business productivity and might not be equally convenient for all the employees. In this regard, it also stated that working from home with the use of recent technologies would only be beneficial for service sector employees but not for industries, which needs physical presence and outputs. Thirdly, voluntary self-isolation and quarantine of contacts are considered effective and acceptable. Also, when combined measures are in place, these can reduce the peak caseload and attack rate as well; however, there is an increased risk of intra-household transmission from index cases to contacts. It also added that those who are isolated and quarantined are likely to undergo distress due to fear and risk perceptions. Lastly, it reported that mobility restrictions are effective only if high travel restrictions are in place, and surprisingly, cancellations of mass events and gatherings have not proven to be effective during the influenza pandemic.

Besides, in a recent epidemic network model by Leung KY et al., demonstrated the potential negative effects of social distancing during an epidemic at the population level [36]. The whole idea of the authors of this study is that rational preventive measures during epidemics could have a counter effect on the population in the long run as these measures might impact the behavioural changes of the public towards others, which could worsen the epidemic outcomes. However, policies and decision-making regarding mitigation strategies should not be generalized depending upon the dynamics of other pandemic modelling studies. Looking at the current role that is being played by the USPs and social distancing measures in combating the spread of COVID-19, it is worth and wise to implement and adhere to such strategies. In regard to this, the UK government has changed its action plan in dealing with the COVID-19 following a modelling study that estimated 260,000 potential deaths [37]. As a result, it decided to implement a combination of social distancing of the entire population, home isolation of infected cases and to quarantine the family members of those who are being infected, and possible school and university closures in order to prevent those huge number of potential deaths. COVID-19 pandemic has taught various lessons to the respective government bodies and healthcare sectors in dealing with this outbreak and how to be prepared in facing such types of outbreaks in the future [38].

Lastly, careful considerations of USPs and mitigating measures should be kept in mind when making pandemic plans, especially aspects such as public compliance and resource planning. Timely implementation of these safety measures in the community would significantly prevent the widespread of the infection. However, future research focussing on the transmission dynamics of the infection would be warranted. There are certain limitations that should be considered when interpreting the findings of the current study. Firstly, as this is a cross-sectional survey, causal inferences cannot be made, and chances for the recall and information bias may exist. Secondly, as the questionnaire is self-administered and thus, by depending upon the self-reported data, it is difficult to predict and understand whether the respondents are filling the survey honestly, i.e., social desirability bias and the responses provided by the participants may not reflect the reality. Lastly, as this is an internet-based online survey, it might not capture the responses from the regions with the restricted access to the social media, and thus may introduce demographic selection bias and might have received the responses mostly from the younger and internet-active population leading to coverage bias.

## Conclusions

In light of the recent pandemic, i.e., COVID-19, although knowledge and beliefs of the general public in India towards USPs are encouraging, there is a need for long term educational interventions as the dynamics and severity of COVID-19 has been keep changing day-by-day rapidly. The findings of this study could guide the public health authorities in making and implementing decisions to combat this pandemic. However, further assessment of factors influencing compliance with such measures is warranted.

## Data Availability

All the information related to the study is embedded within the manuscript.

## Acknowledgments

Authors of this study would like to extend their gratitude towards all the study participants for investing their time, interest and voluntary participation in providing the essential information.

## IRB/IEC approval

This study received an exemption for minimal risk research by the KLE University Institutional Review Board, Karnataka, India on March 2, 2020.

## Authors’ contributions

Study design, development of survey instrument, literature review, drafting the manuscript- SKG; data collection and analysis- MC; reliability & validity of the questionnaire, pilot testing, distribution of the survey questionnaire, proofreading- KU, RV, SN & KKT.

## Available data and materials

All the information related to the study is embedded within the manuscript.

